# Association between Discrimination in Healthcare and Doctor Visits Over Time

**DOI:** 10.1101/2025.04.14.25325800

**Authors:** Michael D. Green, Qing Yang, Hanzhang Xu, Radha Dhingra, Heather R. Farmer, M. Alan Brookhart, Ann Marie Navar, Emily C. O’Brien, Roland J. Thorpe, Matthew E. Dupre

## Abstract

**Objective:** Discrimination in healthcare can disrupt trust. However, it is unknown whether discrimination is associated with the frequency and distribution of healthcare utilization. We assessed how perceived discrimination in healthcare settings is associated with longitudinal patterns of doctor visits among middle-aged and older adults in the United States.

**Study Setting and Design:** Prospective cohort study of US adults who were followed every 2 years for up to 12 years. Discrimination in healthcare was self-reported at baseline (“receiving poorer treatment than others from doctors or hospitals”) and dichotomized (any vs. never). The primary outcome was the patterns of doctor visits over follow-up identified using group-based trajectory models (GBTM).

**Data source:** Health and Retirement Study (HRS) from 2008-2020.

**Principle findings:** Among 13,422 participants (mean age 63.0 years [±8.0]), approximately 19.4%, reported experiencing discrimination in healthcare at baseline. Discrimination in healthcare was more prevalent among participants with more visits, greater disease burden, and greater social vulnerability. Five major patterns of doctor visits over time were identified: “Low-Stable” (∼2 visits/year, 35.5%), “Moderate-Stable” (∼4.5 visits/year, 33.3%), “Increasing” (∼4.5 to 9 visits/year, 13.7%), “Decreasing” (∼13.5 to 6 visits/year, 10.3%), and “Persistently-High” (∼11+ visits/year, 6.3%). Compared to participants with Low-Stable doctor visits, those who reported discrimination were significantly more likely to exhibit Decreasing (relative risk ratio [RRR]=1.38; 95% CI, 1.17–1.63) and Persistently-High (RRR=1.62; 95% CI, 1.30–2.03) patterns of doctor visits.

**Conclusions:** Discrimination in healthcare settings is associated with greater numbers of doctor visits, indicating potential opportunities to improve quality of care.

*What is known on this topic:*

- Perceived discrimination in healthcare is a potential barrier to quality care and is frequently associated with poor health outcomes and patient mistrust in clinical care.

*What this study found:*

- Middle-aged and older adults who perceived discrimination in healthcare settings were significantly more likely to have high-frequency patterns of doctor visits.
- This association persisted after accounting for sociodemographic factors, behavioral factors, and health status.

## Introduction

Experiencing discrimination in healthcare is a barrier to equitable healthcare delivery, and limits the possibility of optimal health outcomes.^1–3^ From a patient standpoint, negative experiences in healthcare can erode trust in providers and the healthcare system,^4^ which in turn, can influence how patients engage with health systems and the management of their health conditions.^3,5,6^ Despite this, the longitudinal association between experiencing discrimination and healthcare utilization is unknown.

Discrimination in healthcare is when people are treated unfairly or overlooked because of who they are or who others think they are.^7^ Reasons that people attribute experiences of discrimination can include sociodemographic characteristics such as age, sex, and race, as well as social class, sexual orientation, or other factors.^8^ Race, ethnicity, and disability are among the factors most commonly associated with unfair treatment in healthcare settings.^9,10^ Patients who experience discrimination in healthcare settings may exhibit distinct patterns in their doctor visits over time;^11^ and these differences could stem from various reasons.^12–15^ For example, discrimination may signal poor quality of care leading to unmet medical needs, worsening health-status, and, as a result, more frequent or fragmented healthcare utilization.^16^ Additionally, individual health needs significantly influence healthcare-seeking behaviors.^17,18^ Middle-aged and older adults have greater healthcare needs.^19,20^ These adults also exhibit a high likelihood of experiencing discrimination in healthcare settings.^21,22^ Therefore, it is crucial to consider both sociodemographic characteristics that are associated with perceived discrimination and markers of health status that need different treatment intensities,^11,23–25^ where treatment plans and the number of visits will differ for conditions with varying etiologies and prognoses.^26^ Identifying heterogenous patterns of healthcare utilization will help us better understand the potential implications of discrimination.

Using the largest ongoing prospective cohort study of middle-aged and older adults in the United States, we examined whether individuals who reported discrimination in healthcare had distinct patterns of doctor visits over time. To achieve this objective, we first identified the major patterns of doctor visits occurring during follow-up. We then assessed how perceived discrimination was associated with the likelihood of exhibiting specific patterns of doctor visits. Finally, the associations were assessed while adjusting for a range of sociodemographic, behavioral, and health-related factors.

## Methods

### Study Participants

Our analysis used data from the Health and Retirement Study (HRS), the largest ongoing nationally representative longitudinal study of U.S. adults over age 50. The HRS is sponsored by the National Institute on Aging (NIA U01AG009740) and is conducted by the University of Michigan.^27^ The HRS has accumulated over three decades of data on more than 40,000 individuals since its launch in 1992. The core survey for the HRS is conducted every 2 years, and comprehensive details on its methodology and response rates are documented elsewhere.^28^ Beginning in 2006, the HRS selected a random half-sample of respondents to collect detailed self-reported psychosocial data.^29,30^ Subsequent psychosocial data were collected from the remaining half-sample in 2008, and follow-up data collection occurred every four years after each half-sample initially provided data through 2020.

The study was limited to adults aged 50-80 at baseline who responded to the question about their exposure to discrimination in healthcare settings from 2008 to 2020 (n=17,642). We examined the patterns of doctor visits for individuals up to age 80 at baseline to limit the influence of healthcare utilization related to end-of-life care at advanced ages. We excluded individuals who did not report at least one doctor visit or hospitalization at baseline (n=1,619) to capture those with potential exposure to discrimination in a healthcare setting. We also excluded individuals who provided only baseline data and lacked longitudinal follow-up (n=2,257). Participants with missing data on study covariates (n=344; ∼2.5%) were also excluded from the analyses. The final analytic sample included 13,422 individuals who provided at least two waves of follow-up data with non-missing doctor visit information over the 14 years of follow-up. All HRS participants provided written informed consent. Our study was approved by the Duke University Health System Institutional Review Board (Pro00108869).

### Discrimination in Healthcare Settings

Perceived discrimination in healthcare settings was measured using an item from the Everyday Discrimination Scale (EDS), a validated instrument designed to capture the frequency of discriminatory experiences across various daily contexts, including interactions in retail, dining, and other social environments.^31^ In 2008, the HRS incorporated an additional item into the psychosocial survey specifically focused on discrimination in healthcare settings, which asked respondents, “In your day-to-day life how often have any of the following things happened to you: You receive poorer service or treatment than other people from doctors or hospitals.” Due to the limited variability in the reported frequency of experiencing healthcare discrimination, we dichotomized the responses to distinguish those who had never encountered discrimination (coded 0) from those who had (coded 1)—which included “less than once a year,” “a few times a year,” “a few times a month,” “at least once a week,” and “almost every day”—an approach consistent with previous research.^32,33^

The primary analyses included self-reported exposure to discrimination in healthcare at baseline. However, we recognize that it is difficult to disentangle the temporality of the association between reported discrimination in healthcare settings and numbers of doctor visits. To further account for this, we conducted additional analyses that included discrimination in health as a time-varying exposure (as detailed below).

### Outcome

Participants were asked at each wave if they had visited a doctor in the past two years and, if so, how many times. Preliminary analyses assessed the full range of reported doctor visits (0-900) and several winsorized thresholds were evaluated for upper limits (i.e., 20+, 30+, 40+, and 50+ visits) to account for extreme/uncommon values. Our final analyses included counts of doctor visits from 0-50+ with approximately 2.92% (n=390) of participants reporting more than 50 doctor visits in a reported wave.

### Covariates

Analyses included the participants’ baseline sociodemographic characteristics, health behaviors, and clinical characteristics to account for factors that could influence exposure to discrimination and/or patterns of healthcare utilization. Sociodemographic factors included age (in years), gender (female or male), self-reported race and ethnicity (Non-Hispanic White, Non-Hispanic Black, or Hispanic), educational attainment (in years), total household wealth based on assets per household member (continuous, log-transformed), marital status (currently married/partnered or not), and health insurance status (insured or uninsured). Health behaviors included smoking status (never smoker, former smoker, or current smoker), daily alcohol consumption (0 drinks, 1-2 drinks, or 3+ drinks), and physical inactivity (physically active or not). Measures of health status included number of difficulties with activities of daily living ([ADL]s; count), body mass index (BMI), and self-reported doctor diagnoses of high blood pressure, diabetes, heart disease, stroke, cancer, and arthritis.

### Statistical Analysis

We use group-based trajectory models (GBTM) to examine underlying heterogeneity in the longitudinal patterns of doctor visits during follow-up. The GBTM were estimated with the *traj* package in Stata 18.5.^34^ This data-driven approach allowed for the identification of distinct subgroups within the population that exhibited similar trajectories of doctor visits over time.^35,36,37^ The optimal number of groups was identified based on assessments of Bayesian information criteria (BIC), average posterior probabilities (AvePP), odds of correct classification (OCC), 95% confidence intervals (CI), and entropy.^38,39^ In our preliminary analyses, we assessed the distributions of doctor visits and determined that zero-inflated Poisson models were most appropriate for modeling the counts of doctor visits over time.

Three sets of sensitivity analyses were also conducted. First, we examined mortality as a potential source of non-random attrition using the “dropout” function in *traj*.^36,40^ Mortality over the 14-year follow-up period was generally low (16.6%) and did not change the results. Second, we evaluated the results using baseline and time-varying indicators of discrimination in our models. Approximately 82% of participants reported the same exposure to discrimination over the entire follow-up period (with about 10% reporting no discrimination at baseline and subsequently reporting discrimination in a later wave). The parameter estimates for the trajectory models with the baseline indicator are available in (Supplementary Table 1). Because there was substantially less follow-up data available for discrimination (measured quadrennially; n=28,673 observations) that would omit nearly half of the doctor visits (measured biennially; n=58,022 observations), our primary analyses included the baseline indicator of discrimination in healthcare. Finally, we included time-varying adjustments for discrimination and major chronic conditions at baseline (i.e. whether an individual had heart disease and/or cancer) to account for potential changes in experiences of discrimination and health status.

Next, we used multinomial logistic regression models to examine the associations between discrimination in healthcare settings reported at baseline and patterns of care visit over time (trajectory group membership), in unadjusted (model 1) and covariate-adjusted (model 2) models. For the multinomial logistic regression models, we used the most common trajectory of doctor visits identified from GBTM as the reference group to compare with other trajectories. We also examined differences among patients with similar starting (or ending) numbers of doctor visits over time. As a final sensitivity analysis, we assessed the multinomial logistic regression models in the trajectory model that included adjustments for time-varying discrimination and comorbidities. *P* values < 0.05 were considered statistically significant. All analyses were performed using Stata 18.5 (StataCorp LP, College Station, TX).

## Results

The average age of our participants at baseline was 63.05 years (standard deviation ± 7.97), and 19.39% reported any discrimination in healthcare. Overall, reports of discrimination in healthcare were significantly higher among adults who were relatively younger, unmarried/unpartnered, uninsured, and belonged to a racial/ethnic minority group (Table 1).

**Table 1.** Baseline Characteristics of by Discrimination Experience, Health and Retirement Study 2008-2020. (n=13,422)

| Variable | No Discrimination,<br>10,836 (80.6%) | Discrimination,<br>2,606 (19.4%) | P-Value |
| --- | --- | --- | --- |
| Age | 63.4 (8.0) | 61.6 (7.7) | <0.001 |
| Male | 4,429 (40.9%) | 1,121 (43.0%) | 0.046 |
| Race/Ethnicity |  |  | <0.001 |
| Non-Hispanic White | 8,027 (74.1%) | 1,771 (68.0%) |  |
| Non-Hispanic Black | 2,007 (18.5%) | 641 (24.6%) |  |
| Hispanic | 802 (7.4%) | 194 (7.4%) |  |
| Married/Partnered | 7,015 (64.7%) | 1,528 (58.6%) | <0.001 |
| Education (Years) | 13.2 (2.8) | 13.2 (2.9) | 0.265 |
| Wealth (Log) | 5.70 (3.44) | 5.17 (3.85) | <0.001 |
| Uninsured | 654 (6.0%) | 230 (8.8%) | <0.001 |
| Smoking Status |  |  | <0.001 |
| Never Smoker | 4,779 (44.1%) | 1,073 (41.2%) |  |
| Former Smoker | 4,450 (41.1%) | 1,063 (40.8%) |  |
| Current Smoker | 1,607 (14.8%) | 470 (18.0%) |  |
| Drinking Status |  |  | <0.001 |
| Moderate | 2,380 (22.0%) | 560 (21.5%) |  |
| None | 6,342 (58.5%) | 1,618 (62.1%) |  |
| Heavy | 2,114 (19.5%) | 428 (16.4%) |  |
| Physically Inactive | 5,564 (51.4%) | 1,446 (55.5%) | <0.001 |
| ADL Limitations | 0.23 (0.78) | 0.45 (1.09) | <0.001 |
| BMI | 29.1 (6.2) | 30.2 (6.7) | <0.001 |
| Hypertension | 5,979 (55.2%) | 1,503 (57.7%) | 0.021 |
| Heart Problem | 1,986 (18.3%) | 518 (19.9%) | 0.068 |
| Stroke | 640 (5.9%) | 187 (7.2%) | 0.015 |
| Diabetes | 2,110 (19.5%) | 611 (23.5%) | <0.001 |
| Cancer | 1,248 (11.5%) | 333 (12.8%) | 0.073 |
| Arthritis | 5,740 (53.0%) | 1,492 (57.3%) | <0.001 |
| Psychiatric Condition | 1,652 (15.3%) | 629 (24.1%) | <0.001 |
| Note: Parenthesis next to continuous variables are standard deviations. P-values generated from Kruskal-Wallis tests and Chi-Square Tests. Abbreviations: BMI= Body Mass Index, ADL=Activities of Daily Living |  |  |  |

Participants who reported discrimination were also more likely to have higher BMI, more functional limitations, and higher prevalence of chronic conditions.

### Patterns of Doctor Visits Over Time

We identified five distinct patterns of doctor visits over time (Figure 1). Baseline characteristics of individuals across the five trajectory groups are presented in Table 2. The two most common patterns of doctor visits were relatively stable over time, representing approximately 2 yearly visits (“Low-Stable” group; 36.51%) and approximately 4.5 yearly doctor visits (“Moderate-Stable” group; 33.26%). Two additional patterns showed significant changes in doctor visits over time. The “Increasing” group (13.70%) indicated about 4.5 doctor visits each year and gradually increased to more than 9 yearly doctor visits over the study period.

**Figure 1.**
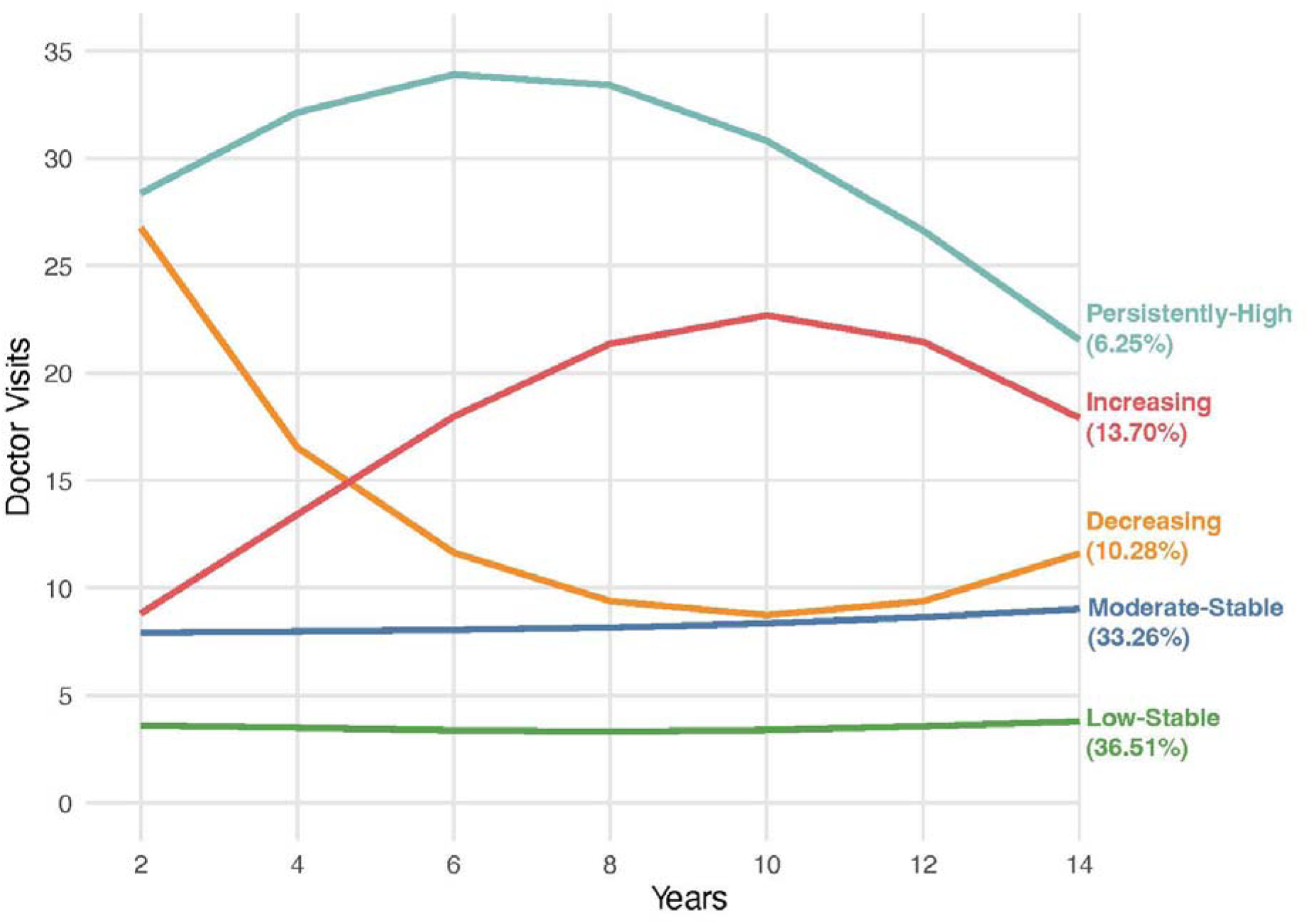
Estimated Trajectories of Doctor Visits Over Time in Middle-Aged and Older Adults, Health and Retirement Study 2008-2020 (N=13,422). *Note*: Estimates were derived using a group-based trajectory model that assumed a zero-inflated Poisson distribution for the number of doctor visits over time.

**Table 2.** Baseline Characteristics of Individuals in Identified Trajectory Groups of Doctor Visits, Health and Retirement Study 2008-2020. (n=13,422)

| Variables | Trajectory of Doctor Visits |  |  |  |  |
| --- | --- | --- | --- | --- | --- |
|  | Low-Stable | Moderate-Stable | Decreasing | Increasing | Persistently-High |
| n | 4940 (36.51%) | 4512 (33.26%) | 1392 (10.28%) | 1773 (13.70%) | 825 (6.25%) |
| Discrimination in Healthcare | 842 (17.0%) | 851 (18.9%) | 334 (24.0%) | 335 (18.9%) | 244 (29.6%) |
| Age | 61.55 (7.88) | 63.72 (7.83) | 64.42 (7.93) | 64.19 (8.00) | 63.59 (7.97) |
| Male Gender | 2250 (45.5%) | 1776 (39.4%) | 529 (38.0%) | 705 (39.8%) | 290 (35.2%) |
| Race and Ethnicity |  |  |  |  |  |
| Non-Hispanic White | 3480 (70.4%) | 3377 (74.8%) | 984 (70.7%) | 1329 (75.0%) | 628 (76.1%) |
| Non-Hispanic Black | 1022 (20.7%) | 852 (18.9%) | 295 (21.2%) | 321 (18.1%) | 158 (19.2%) |
| Hispanic | 438 (8.9%) | 283 (6.3%) | 113 (8.1%) | 123 (6.9%) | 39 (4.7%) |
| Married/Partnered | 3218 (65.1%) | 2933 (65.0%) | 807 (58.0%) | 1093 (61.6%) | 492 (59.6%) |
| Education, Years | 13.19 (2.83) | 13.36 (2.72) | 12.96 (2.83) | 13.16 (2.84) | 13.08 (2.74) |
| Wealth | 5.64 (3.23) | 5.80 (3.45) | 5.28 (3.98) | 5.43 (3.75) | 5.09 (4.18) |
| Uninsured | 434 (8.8%) | 267 (5.9%) | 54 (3.9%) | 97 (5.5%) | 32 (3.9%) |
| Smoking Status |  |  |  |  |  |
| Never Smoker | 2287 (46.3%) | 1997 (44.3%) | 547 (39.3%) | 700 (39.5%) | 321 (38.9%) |
| Former Smoker | 1846 (37.4%) | 1914 (42.4%) | 620 (44.5%) | 780 (44.0%) | 353 (42.8%) |
| Current Smoker | 807 (16.3%) | 601 (13.3%) | 225 (16.2%) | 293 (16.5%) | 151 (18.3%) |
| Drinking Status |  |  |  |  |  |
| Moderate Consumption (1-2 drinks daily) | 1266 (25.6%) | 1019 (22.6%) | 225 (16.2%) | 324 (18.3%) | 106 (12.8%) |
| No Consumption (0 drinks daily) | 2641 (53.5%) | 2633 (58.4%) | 956 (68.7%) | 1134 (64.0%) | 596 (72.2%) |
| Heavy Consumption (3+ drinks daily) | 1033 (20.9%) | 860 (19.1%) | 211 (15.2%) | 315 (17.8%) | 123 (14.9%) |
| Physically Inactive | 2155 (43.6%) | 2322 (51.5%) | 921 (66.2%) | 1040 (58.7%) | 572 (69.3%) |
| ADL Limitations | 0.07 (0.26) | 0.12 (0.32) | 0.25 (0.44) | 0.18 (0.38) | 0.34 (0.47) |
| Body Mass Index | 28.42 (5.67) | 29.56 (6.30) | 29.85 (6.79) | 29.84 (6.50) | 30.97 (7.62) |
| Hypertension | 2151 (43.5%) | 2704 (59.9%) | 941 (67.6%) | 1130 (63.7%) | 556 (67.4%) |
| Heart Problem | 534 (10.8%) | 849 (18.8%) | 401 (28.8%) | 428 (24.1%) | 292 (35.4%) |
| Stroke | 183 (3.7%) | 252 (5.6%) | 139 (10.0%) | 137 (7.7%) | 116 (14.1%) |
| Diabetes | 551 (11.2%) | 1013 (22.5%) | 402 (28.9%) | 482 (27.2%) | 273 (33.1%) |
| Cancer | 371 (7.5%) | 527 (11.7%) | 264 (19.0%) | 256 (14.4%) | 163 (19.8%) |
| Arthritis | 1950 (39.5%) | 2554 (56.6%) | 942 (67.7%) | 1155 (65.1%) | 631 (76.5%) |
| Psychiatric Condition | 468 (9.5%) | 766 (17.0%) | 324 (23.3%) | 417 (23.5%) | 306 (37.1%) |

Continuous variables presented as means with standard deviations in parenthesis. Categorical and binary variables presented as counts with percentages in parenthesis. Wealth is log-adjusted. *Abbreviations:* ADL = Activities of Daily Living.

**Table 2.**
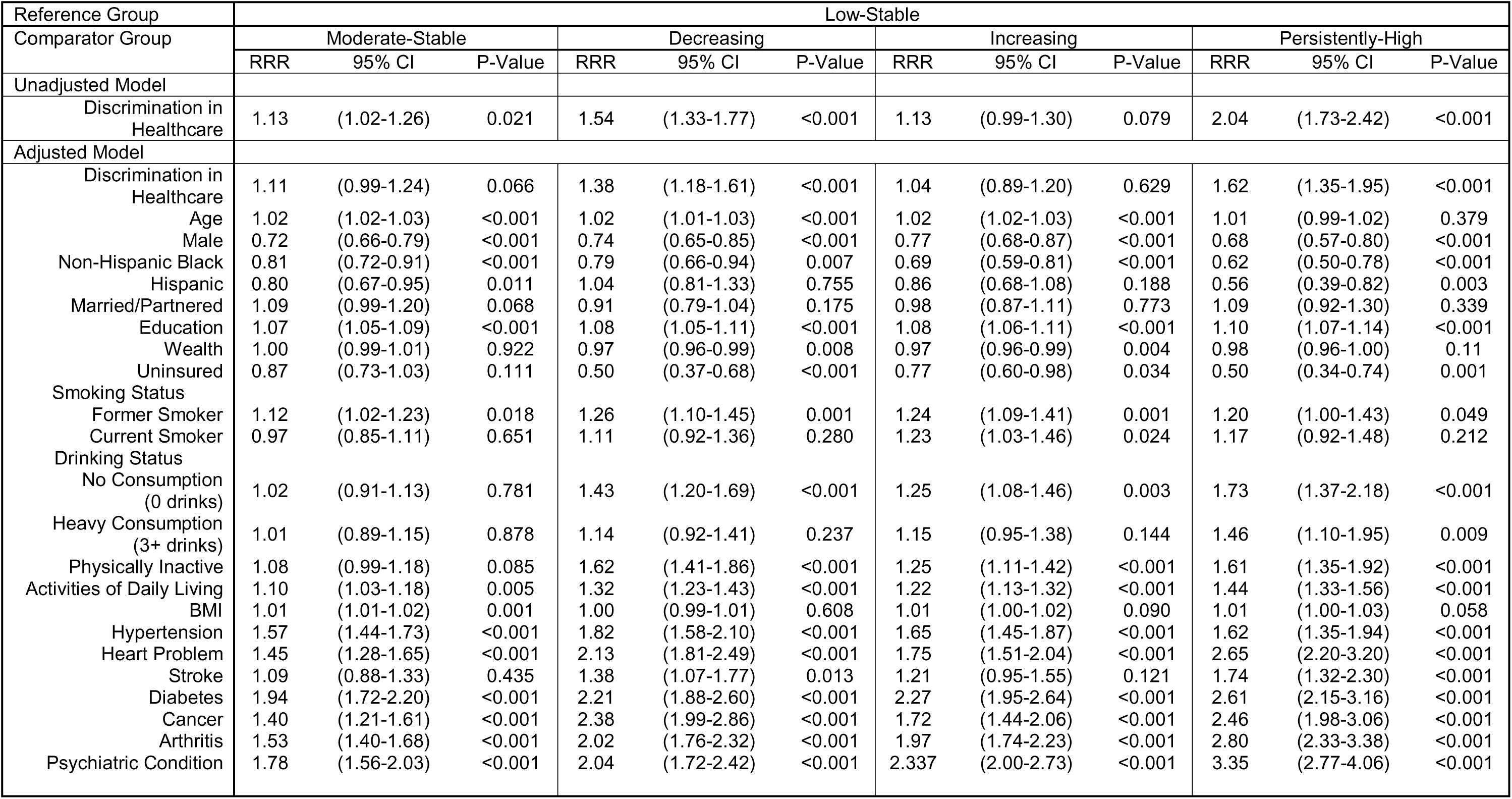
Multinomial Estimates of Association Between Discrimination in Healthcare and Trajectory Group Membership of Middle Aged and Older Adults, Health and Retirement Study 2008-2020. (n=13,422)

| Reference Group | Low-Stable |  |  |  |  |  |  |  |  |  |  |  |
| --- | --- | --- | --- | --- | --- | --- | --- | --- | --- | --- | --- | --- |
| Comparator Group | Moderate-Stable |  |  | Decreasing |  |  | Increasing |  |  | Persistently-High |  |  |
|  | RRR | 95% CI | P-Value | RRR | 95% CI | P-Value | RRR | 95% CI | P-Value | RRR | 95% CI | P-Value |
| Unadjusted Model |  |  |  |  |  |  |  |  |  |  |  |  |
| Discrimination in Healthcare | 1.13 | (1.02-1.26) | 0.021 | 1.54 | (1.33-1.77) | <0.001 | 1.13 | (0.99-1.30) | 0.079 | 2.04 | (1.73-2.42) | <0.001 |
| Adjusted Model |  |  |  |  |  |  |  |  |  |  |  |  |
| Discrimination in Healthcare | 1.11 | (0.99-1.24) | 0.066 | 1.38 | (1.18-1.61) | <0.001 | 1.04 | (0.89-1.20) | 0.629 | 1.62 | (1.35-1.95) | <0.001 |
| Age | 1.02 | (1.02-1.03) | <0.001 | 1.02 | (1.01-1.03) | <0.001 | 1.02 | (1.02-1.03) | <0.001 | 1.01 | (0.99-1.02) | 0.379 |
| Male | 0.72 | (0.66-0.79) | <0.001 | 0.74 | (0.65-0.85) | <0.001 | 0.77 | (0.68-0.87) | <0.001 | 0.68 | (0.57-0.80) | <0.001 |
| Non-Hispanic Black | 0.81 | (0.72-0.91) | <0.001 | 0.79 | (0.66-0.94) | 0.007 | 0.69 | (0.59-0.81) | <0.001 | 0.62 | (0.50-0.78) | <0.001 |
| Hispanic | 0.80 | (0.67-0.95) | 0.011 | 1.04 | (0.81-1.33) | 0.755 | 0.86 | (0.68-1.08) | 0.188 | 0.56 | (0.39-0.82) | 0.003 |
| Married/Partnered | 1.09 | (0.99-1.20) | 0.068 | 0.91 | (0.79-1.04) | 0.175 | 0.98 | (0.87-1.11) | 0.773 | 1.09 | (0.92-1.30) | 0.339 |
| Education | 1.07 | (1.05-1.09) | <0.001 | 1.08 | (1.05-1.11) | <0.001 | 1.08 | (1.06-1.11) | <0.001 | 1.10 | (1.07-1.14) | <0.001 |
| Wealth | 1.00 | (0.99-1.01) | 0.922 | 0.97 | (0.96-0.99) | 0.008 | 0.97 | (0.96-0.99) | 0.004 | 0.98 | (0.96-1.00) | 0.11 |
| Uninsured | 0.87 | (0.73-1.03) | 0.111 | 0.50 | (0.37-0.68) | <0.001 | 0.77 | (0.60-0.98) | 0.034 | 0.50 | (0.34-0.74) | 0.001 |
| Smoking Status |  |  |  |  |  |  |  |  |  |  |  |  |
| Former Smoker | 1.12 | (1.02-1.23) | 0.018 | 1.26 | (1.10-1.45) | 0.001 | 1.24 | (1.09-1.41) | 0.001 | 1.20 | (1.00-1.43) | 0.049 |
| Current Smoker | 0.97 | (0.85-1.11) | 0.651 | 1.11 | (0.92-1.36) | 0.280 | 1.23 | (1.03-1.46) | 0.024 | 1.17 | (0.92-1.48) | 0.212 |
| Drinking Status |  |  |  |  |  |  |  |  |  |  |  |  |
| No Consumption (0 drinks) | 1.02 | (0.91-1.13) | 0.781 | 1.43 | (1.20-1.69) | <0.001 | 1.25 | (1.08-1.46) | 0.003 | 1.73 | (1.37-2.18) | <0.001 |
| Heavy Consumption (3+ drinks) | 1.01 | (0.89-1.15) | 0.878 | 1.14 | (0.92-1.41) | 0.237 | 1.15 | (0.95-1.38) | 0.144 | 1.46 | (1.10-1.95) | 0.009 |
| Physically Inactive | 1.08 | (0.99-1.18) | 0.085 | 1.62 | (1.41-1.86) | <0.001 | 1.25 | (1.11-1.42) | <0.001 | 1.61 | (1.35-1.92) | <0.001 |
| Activities of Daily Living | 1.10 | (1.03-1.18) | 0.005 | 1.32 | (1.23-1.43) | <0.001 | 1.22 | (1.13-1.32) | <0.001 | 1.44 | (1.33-1.56) | <0.001 |
| BMI | 1.01 | (1.01-1.02) | 0.001 | 1.00 | (0.99-1.01) | 0.608 | 1.01 | (1.00-1.02) | 0.090 | 1.01 | (1.00-1.03) | 0.058 |
| Hypertension | 1.57 | (1.44-1.73) | <0.001 | 1.82 | (1.58-2.10) | <0.001 | 1.65 | (1.45-1.87) | <0.001 | 1.62 | (1.35-1.94) | <0.001 |
| Heart Problem | 1.45 | (1.28-1.65) | <0.001 | 2.13 | (1.81-2.49) | <0.001 | 1.75 | (1.51-2.04) | <0.001 | 2.65 | (2.20-3.20) | <0.001 |
| Stroke | 1.09 | (0.88-1.33) | 0.435 | 1.38 | (1.07-1.77) | 0.013 | 1.21 | (0.95-1.55) | 0.121 | 1.74 | (1.32-2.30) | <0.001 |
| Diabetes | 1.94 | (1.72-2.20) | <0.001 | 2.21 | (1.88-2.60) | <0.001 | 2.27 | (1.95-2.64) | <0.001 | 2.61 | (2.15-3.16) | <0.001 |
| Cancer | 1.40 | (1.21-1.61) | <0.001 | 2.38 | (1.99-2.86) | <0.001 | 1.72 | (1.44-2.06) | <0.001 | 2.46 | (1.98-3.06) | <0.001 |
| Arthritis | 1.53 | (1.40-1.68) | <0.001 | 2.02 | (1.76-2.32) | <0.001 | 1.97 | (1.74-2.23) | <0.001 | 2.80 | (2.33-3.38) | <0.001 |
| Psychiatric Condition | 1.78 | (1.56-2.03) | <0.001 | 2.04 | (1.72-2.42) | <0.001 | 2.337 | (2.00-2.73) | <0.001 | 3.35 | (2.77-4.06) | <0.001 |

Conversely, the “Decreasing” group (10.28%) showed an inverse pattern, starting with over 13 yearly visits and gradually decreasing to a lower but still frequent level of about 6 yearly visits. Lastly, there was a “Persistently-High” group (6.25%) which characterized participants with consistently high numbers of doctor visits throughout the study period, with nearly monthly visits.

Across all groups, the majority of individuals were Non-Hispanic White. The Low-Stable group was the youngest (mean age 61.6), with the lowest rates of most chronic health conditions and lowest levels of discrimination in healthcare (17.0%). The Moderate-Stable group had relatively low experiences of discrimination (18.9%) but a greater burden of chronic conditions. The Decreasing group reported greater discrimination in healthcare (24.0%) was the oldest on average (64.4 years) and faced significant health challenges. The Increasing group (13.7%) had a similar health profile to the Decreasing group, with a high prevalence of chronic conditions but reported moderate discrimination (18.9%). The Persistently-High group (6.3%) had the poorest health. This group had the lowest socioeconomic status and the highest reported discrimination in healthcare (29.6%).

### Association between Discrimination and Trajectories

Table 3 presents results from the multinomial logistic regression models of the association between discrimination in healthcare and doctor visit trajectory group membership. Overall, in the unadjusted models, individuals who reported discrimination in healthcare settings had significantly higher likelihood of following trajectories with more frequent doctor visits than those who did not report discrimination. Compared with the Low-Stable group, results showed that discrimination was associated with a 13% higher relative risk of being in the Moderate-Stable group (relative risk ratio [RRR]=1.13, *P*=0.021), a 54% higher relative risk of being in the Decreasing group (RRR=1.54, *P*<0.001), and more than double the relative risk of being in the Persistently-High group (RRR=2.04, *P*<0.001). After adjusting for sociodemographic, behavioral, and health-related covariates, the associations were attenuated but remained significant for the Decreasing (RRR=1.38, *P*<0.001) and Persistently-High (RRR=1.62, *P*<0.001) groups only. The association with the Moderate-Stable group was no longer statistically significant (RRR=1.11, *P*=0.066).

**Table 3.**
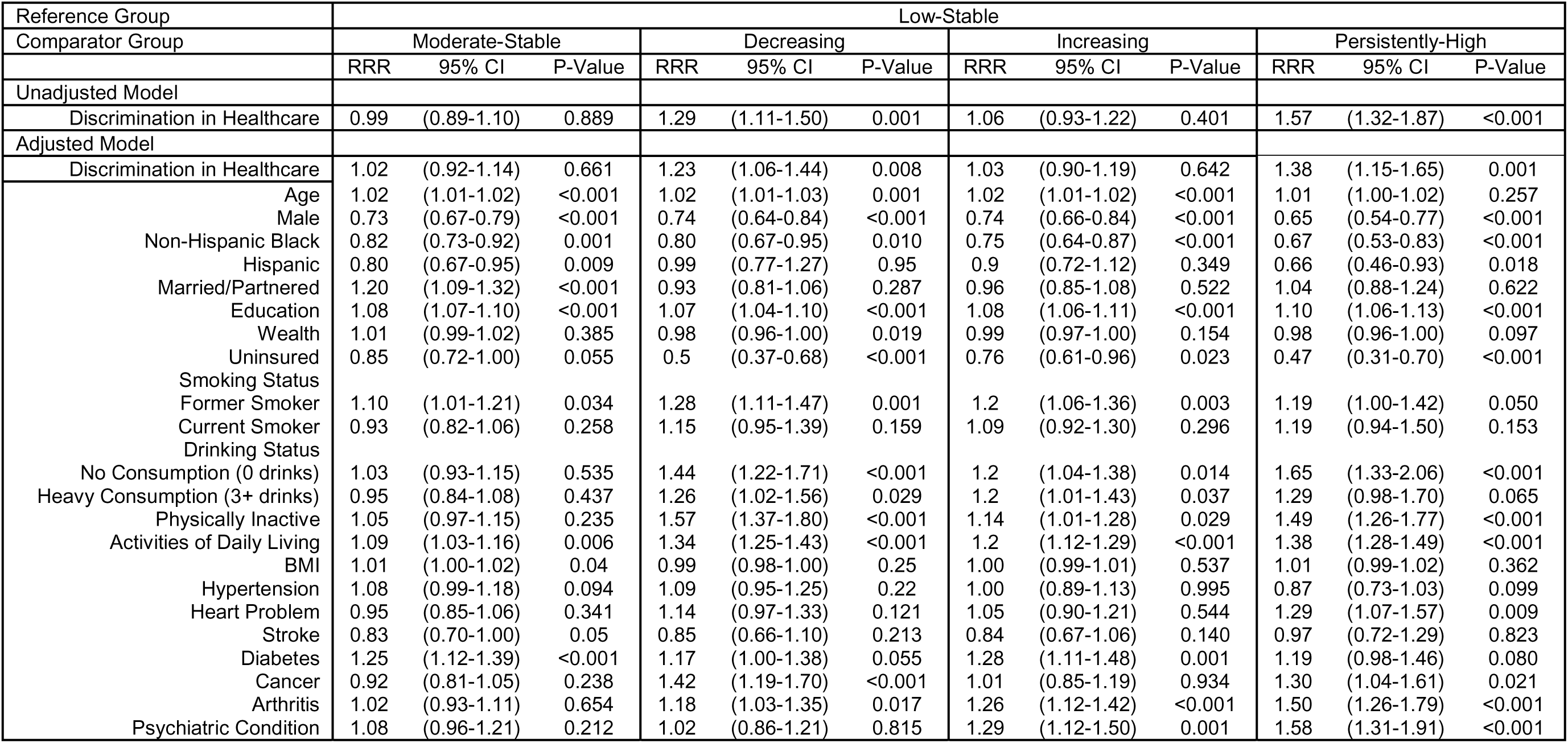
Multinomial Estimates of Association Between Discrimination in Healthcare and Trajectory Group Membership of Middle Aged and Older Adults, Health and Retirement Study 2008-2020. (n=13,422)

Supplementary Figure 1 illustrates the results from the multinomial logistic regression models using different reference groups that reflect similar levels of doctor visits at the start or end of the trajectory (the estimates are provided in Supplementary Table 2). We highlight these groups to better understand the association between discrimination and changing patterns of doctor visits among individuals who begin or conclude care at comparable levels. When comparing the Decreasing (∼13.5 yearly visits decreasing to ∼6) and the Moderate-Stable (∼4 yearly visits), discrimination was associated with a 24% higher relative risk of membership in the group which starts with high amounts of visits then decreasing. Discrimination was associated with a 36% lower relative risk of starting with a high number of visits in the Increasing (∼4.5 yearly visits increasing to ∼9) compared to the Persistently-High Group (11+ Visits). Finally, when considering time-varying changes in discrimination and major chronic conditions when identifying trajectories of doctor visits (Supplementary Figure 2), the results for discrimination were largely consistent and only partially attenuated (Supplementary Table 3 and Supplementary Table 4).

## Discussion

We identified 5 major longitudinal patterns of doctor visits in a large cohort of U.S. middle-aged and older adults. Our findings suggest that discrimination in healthcare was associated with exhibiting high-frequency patterns doctor visits over time. The associations persisted after accounting for sociodemographic, behavioral, and health factors. Persistent discrimination can hinder effective patient-provider communication, resulting in fragmented or ineffective care that necessitates repeated visits.^41,42^ Additionally, this pattern may reflect a cycle where experiencing discrimination contributes to psychological stress and poor health management, leading to more frequent doctor visits as health conditions progress. This association indicates the potential for discrimination to deepen health inequities, as affected individuals may experience both the direct harm of biased care and the indirect consequences of unmanaged or poorly managed chronic conditions.

We found that middle-aged and older adults who had initially higher numbers of doctor visits declined over long-term follow-up, with varying rates of decline. Discrimination in healthcare was associated with distinct patterns of doctor visits over time. Those who reported discrimination had a significantly higher probability of exhibiting patterns of doctor visits that were initially high, but then declined int frequency (Decreasing) compared to those with consistently low (Low-Stable) or moderate (Moderate-Stable) numbers of visits. Before adjustment for sociodemographic and health factors, discrimination was also associated with a greater probability of demonstrating high numbers of doctor visits (Persistently-High) compared with those who started high and then largely declined (Decreasing). Because the Persistently-High pattern of doctor visits was characterized by worse health status, it is plausible that persistently high doctor visits may be more closely related to clinical needs than to discrimination exposure. In comparison, the pattern of initially high but declining visits appears more distinctively associated with experiencing discrimination in the healthcare setting.

However, because of the difficulty disentangling these factors, these conclusions warrant further investigation.

One concern regarding discrimination in healthcare is that it may lead patients to avoid care entirely. Our findings do not support this interpretation among middle-aged and older adults. Individuals who reported discrimination were more likely to exhibit higher-frequency patterns of visits rather than lower ones. However, a higher volume of visits does not necessarily reflect effective care. Higher frequency of doctor visits among those who report discrimination may reflect poorer quality care rather than higher quality treatment. It is also important to acknowledge that the direction of this association may partly reflect exposure opportunity. Individuals with more doctor visits have more encounters in which discrimination can occur. Those with the greatest clinical need, and therefore the most in contact with the healthcare system, may paradoxically be the most likely to encounter mistreatment. Discrimination in healthcare, as measured in this study, represents a departure from the broader framing of discrimination as a chronic psychological stressor experienced across various settings.^43–45^ By assessing reports of discrimination in healthcare specifically, these findings illuminate potentially actionable targets for health systems seeking to improve care quality.

Assessing discrimination in healthcare can be effectively used as a tool to assess the quality of care the services provided. Studies have shown that the cost of services provided to individuals is higher for those who experience discrimination than those who do not.^46,47^ Quality of care is in the interest of both health systems focused on cost savings, alongside the doctors who seek to improve their patients’ health. Both patterns that exhibited the highest initial volume of doctor visits (Decreasing and Persistently-High) demonstrated declining visits over the follow-up period. Several interpretations of this general pattern are possible. Improvement in health status could reduce the need for visits. For example, early frequent visits could reflect successful management of a new diagnosis or recovery from an acute condition. The decline could also reflect disengagement from care, potentially reacting to accumulated negative healthcare experiences including discrimination, even as clinical need persists. Importantly, even after such declines, these groups maintained frequent visits more so than other groups.

Further research is needed to help clarify whether the declines in visits reflect a change in their health status, or whether discrimination is a driver of this change. Additionally, inclusion of more racial/ethnic groups, consideration of broader age group categories, and time-varying covariates would complement this analysis.

We believe our results are statistically rigorous and present a thoughtful analysis that reflects real-world disease prognosis. GBTM has been critiqued as a method which can generate spurious findings when Average Posterior Probability is the sole criterion for evaluating model fit.^48^ To address this concern we, assessed model fit using Average Posterior Probability, entropy, Bayesian Information Criterion, Odds of Correct Classification, and 95% CIs. We further evaluated our trajectories qualitatively by comparing our generated trajectories to plausible care patterns. For example, if an individual had an onset of a condition such as kidney failure, it would be plausible for them to start out with frequent visits and then require more doctor visits over time. If an individual had a condition that was temporary, it would make sense for them to start out with a high number of doctor visits, then decline.

### Limitations

There are some limitations to consider when interpreting our results. We used a single item to evaluate discrimination, instead of a full-scale assessment. While many scales have been validated for population subgroups, with the intention of better understanding their experiences, few have been constructed for broad use as a quality improvement tool.^49–54^ A psychometrically validated assessment, complemented by qualitative inquiry, could provide additional context for impact of the perceived discrimination on subsequent doctor visits patterns. Our sample is limited to those who responded to the HRS. While this study design and sampling approach are robust, they may have excluded perspectives of marginalized individuals or those who were unable to complete the survey. Furthermore, our approach may not have adequately accounted for unobserved confounding, limiting our ability to draw causal conclusions from our findings. We did not stratify by sociodemographic characteristics for our analysis, which could cause different trends to emerge. For example, individuals who attribute perceived discrimination to a disability are less likely to seek follow-up care.^55^ Evaluating how discrimination impacts the care trajectories of aging populations with disabilities and functional limitations could be a crucial future consideration alongside other populations. The self-perception of aging is both positively and negatively connected to perceived healthcare discrimination.^56^ We did not explore positive perception of aging in our analysis or ageism, but future studies could consider this as a reason for divergence in discrimination experiences and care-seeking behavior amongst middle-aged and older adults.

## Conclusion

Our findings underscore the multifaceted impact of discrimination in healthcare on patterns in healthcare utilization over time. This study highlights the persistent and potentially costly implications of discriminatory experiences. It calls attention to the need for more nuanced, inclusive research that captures the diversity of lived experiences across sociodemographic lines. Addressing discrimination in healthcare is not only a moral imperative, but also a pragmatic one. Discrimination in healthcare directly affects patient trust, care efficiency, and system-wide costs. Interventions designed to reduce discriminatory practices could mitigate the cycle of fragmented care and improve outcomes for those most affected.

## Data Availability

All data produced are available online at rand.org.

## Notes

Funding: The research was supported by a National Institute on Aging (NIA) grant (PI: Dupre, R01AG069938), a NIA Diversity Supplement Award (PI: Dupre, R01AG069938-02S1), and a NIA pre-doctoral fellowship (PI: Green, F99AG088695). The content is solely the responsibility of the authors and does not necessarily represent the official views of the National Institutes of Health.

### Competing Interest Statement

The authors have declared no competing interest.

### Funding Statement

The research was supported by a National Institute on Aging (NIA) grant (PI: Dupre, R01AG069938), a NIA Diversity Supplement Award (PI: Dupre, R01AG069938-02S1), and a NIA pre-doctoral fellowship (PI: Green, F99AG088695). The content is solely the responsibility of the authors and does not necessarily represent the official views of the National Institutes of Health

### Author Declarations

Duke University Health System Institutional Review Board (Pro00108869)

### Summary of Updates

Title change, added additional authors who contributed feedback/revisions on the methods and analysis. Incorporated feedback to relax and implication of causality, alongside consideration of time-varying discrimination and chronic conditions.

