## Supplemental Figures and Tables for "Association between Discrimination in Healthcare and Doctor Visits Over Time"

**Supplementary Table 1.** Parameter Estimates from the Zero-Inflated Poisson Group Based Trajectory Model Characterizing the Trajectory of Doctor Visits Over Time for Middle Aged and Older Adults. (n=13,442)

| Group | Parameter | Estimate | Standard Error | T Score | P Value |
| --- | --- | --- | --- | --- | --- |
| Low-Stable | Parameter for Polynomial (Count) | | | | |
|  | Intercept | 1.332 | 0.008 | 165.424 | <0.001 |
|  | Linear | 0.031 | 0.006 | 5.005 | <0.001 |
|  | Quadratic | 0.015 | 0.006 | 2.493 | 0.0127 |
|  | Parameter for Polynomial (Zero-Inflation) | | | | |
|  | Alpha0 | -2.062 | 0.042 | -49.61 | <0.001 |
|  | Alpha1 | 0.262 | 0.050 | 5.295 | <0.001 |
|  | Alpha2 | -0.596 | 0.052 | -11.422 | <0.001 |
| Moderate-Stable | Parameter for Polynomial (Count) | | | | |
|  | Intercept | 2.135 | 0.007 | 317.055 | <0.001 |
|  | Linear | 0.049 | 0.004 | 11.010 | <0.001 |
|  | Quadratic | 0.007 | 0.005 | 1.422 | 0.1551 |
|  | Parameter for Polynomial (Zero-Inflation) | | | | |
|  | Alpha0 | -3.274 | 0.072 | -45.282 | <0.001 |
|  | Alpha1 | 0.361 | 0.071 | 5.069 | <0.001 |
|  | Alpha2 | -0.404 | 0.077 | -5.235 | <0.001 |
| Decreasing | Parameter for Polynomial (Count) | | | | |
|  | Intercept | 2.298 | 0.015 | 153.664 | <0.001 |
|  | Linear | -0.270 | 0.009 | -30.835 | <0.001 |
|  | Quadratic | 0.266 | 0.011 | 25.1 | <0.001 |
|  | Parameter for Polynomial (Zero-Inflation) | | | | |
|  | Alpha0 | -2.794 | 0.010 | -27.965 | <0.001 |
|  | Alpha1 | 0.331 | 0.102 | 3.249 | 0.0012 |
|  | Alpha2 | -0.394 | 0.112 | -3.515 | 0.0004 |
| Increasing | Parameter for Polynomial (Count) | | | | |
|  | Intercept | 3.112 | 0.007 | 418.683 | <0.001 |
|  | Linear | 0.238 | 0.007 | 34.016 | <0.001 |
|  | Quadratic | -0.252 | 0.006 | -41.959 | <0.001 |
|  | Parameter for Polynomial (Zero-Inflation) | | | | |
|  | Alpha0 | -2.966 | 0.01 | -29.764 | <0.001 |
|  | Alpha1 | 0.136 | 0.117 | 1.168 | 0.2428 |
|  | Alpha2 | -0.598 | 0.122 | -4.922 | <0.001 |
| Persistently-High | Parameter for Polynomial (Count) | | | | |
|  | Intercept | 3.530 | 0.007 | 506.656 | <0.001 |
|  | Linear | -0.089 | 0.008 | -11.141 | <0.001 |
|  | Quadratic | -0.139 | 0.007 | -19.524 | <0.001 |
|  | Parameter for Polynomial (Zero-Inflation) | | | | |
|  | Alpha0 | -3.837 | 0.230 | -16.703 | <0.001 |
|  | Alpha1 | 0.332 | 0.247 | 1.341 | 0.1799 |
|  | Alpha2 | -0.449 | 0.253 | -1.775 | 0.0759 |
| *Note*: Average Posterior Probability for each group were: Low-Stable (0.940), Moderate-Stable (0.897), Decreasing (0.897), Increasing (0.926), Persistently-High (0.970). Model included 58,022 observations. | | | | | |

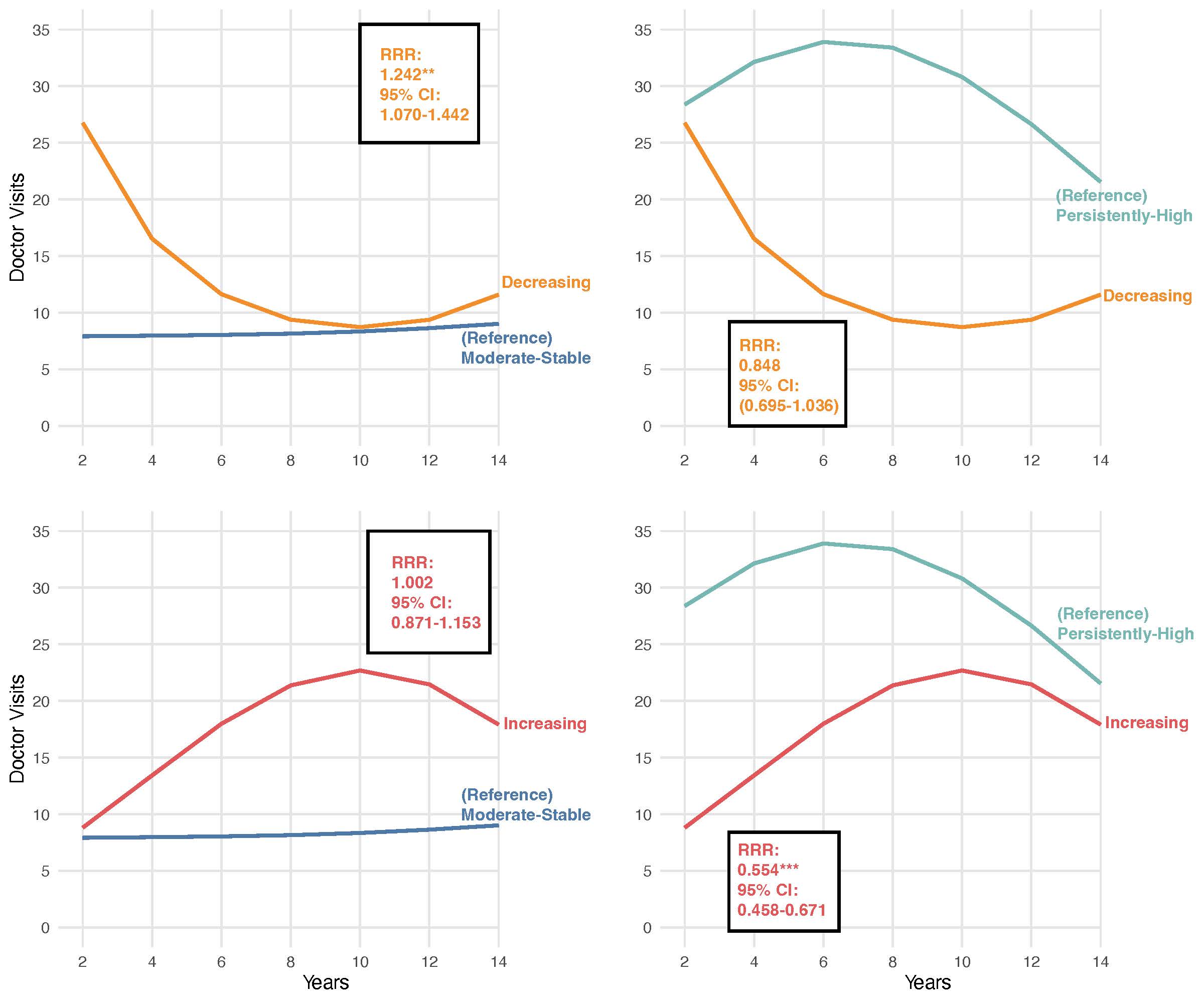

**Supplementary Figure 1.** Multinomial Estimates of Association between Discrimination in Healthcare Settings and Trajectory Group Membership with varying Comparator Groups based on the Similar Number of Doctor Visits, Health and Retirement Study (N=13,422) *Note*: Models adjust for sociodemographic background, health behaviors, and general health status. Reference categories are indicated with parenthesis. **=p<0.01 ***=p<0.001. Abbreviations: RRR = Relative Risk Ratio.

**Supplementary Table 2.** Multinomial Estimates of Association between Discrimination in Healthcare Settings and Trajectory Group Membership with varying Comparator Groups based on the Similar Number of Doctor Visits, Health and Retirement Study (n=13,422)

| Trajectory Group | Decreasing | | | Increasing | | | Decreasing | | | Increasing | | |
| --- | --- | --- | --- | --- | --- | --- | --- | --- | --- | --- | --- | --- |
| (Reference Group) | (Moderate-Stable) | | | (Moderate-Stable) | | | (Persistently-High) | | | (Persistently-High) | | |
|  | RRR | 95% CI | P-Value | RRR | 95% CI | P-Value | RRR | 95% CI | P-Value | RRR | 95% CI | P-Value |
| Unadjusted Model |  |  |  |  |  |  |  |  |  |  |  |  |
| Discrimination in Healthcare | 1.36 | (1.18-1.57) | <0.001 | 1.00 | (0.87-1.15) | 0.975 | 0.75 | (0.62-0.91) | 0.004 | 0.55 | (0.46-0.67) | <0.001 |
| Adjusted Model |  |  |  |  |  |  |  |  |  |  |  |  |
| Discrimination in Healthcare | 1.24 | (1.07-1.44) | 0.004 | 0.935 | (0.81-1.08) | 0.359 | 0.85 | (0.70-1.04) | 0.106 | 0.64 | (0.52-0.78) | <0.001 |
| Age | 1.00 | (0.99-1.01) | 0.368 | 1.00 | (0.99-1.01) | 0.756 | 1.01 | (1.00-1.03) | 0.054 | 1.02 | (1.01-1.03) | 0.004 |
| Male | 1.03 | (0.90-1.18) | 0.706 | 1.07 | (0.94-1.20) | 0.311 | 1.10 | (0.90-1.33) | 0.349 | 1.14 | (0.94-1.37) | 0.175 |
| Non-Hispanic Black | 0.97 | (0.82-1.15) | 0.756 | 0.85 | (0.73-1.00) | 0.048 | 1.26 | (0.99-1.61) | 0.058 | 1.11 | (0.87-1.40) | 0.405 |
| Hispanic | 1.30 | (1.02-1.67) | 0.037 | 1.07 | (0.85-1.36) | 0.569 | 1.85 | (1.24-2.75) | 0.003 | 1.52 | (1.02-2.25) | 0.038 |
| Married | 1.01 | (0.98-1.03) | 0.493 | 1.01 | (0.99-1.04) | 0.302 | 0.98 | (0.94-1.01) | 0.190 | 0.98 | (0.95-1.01) | 0.250 |
| Education | 0.98 | (0.96-0.99) | 0.007 | 0.98 | (0.96-0.99) | 0.004 | 0.99 | (0.97-1.02) | 0.591 | 0.99 | (0.97-1.02) | 0.589 |
| Wealth | 1.13 | (0.98-1.29) | 0.091 | 1.11 | (0.98-1.26) | 0.099 | 1.06 | (0.87-1.28) | 0.596 | 1.04 | (0.86-1.26) | 0.683 |
| Uninsured | 1.15 | (0.95-1.40) | 0.162 | 1.26 | (1.06-1.51) | 0.009 | 0.96 | (0.73-1.25) | 0.740 | 1.05 | (0.81-1.36) | 0.707 |
| Smoking Status |  |  |  |  |  |  |  |  |  |  |  |  |
| Former Smoker | 1.41 | (1.19-1.66) | <0.001 | 1.24 | (1.07-1.43) | 0.005 | 0.83 | (0.64-1.07) | 0.147 | 0.73 | (0.57-0.93) | 0.011 |
| Current Smoker | 1.13 | (0.91-1.40) | 0.272 | 1.14 | (0.95-1.36) | 0.174 | 0.78 | (0.56-1.08) | 0.133 | 0.79 | (0.58-1.07) | 0.124 |
| Drinking Status |  |  |  |  |  |  |  |  |  |  |  |  |
| No Consumption  (0 drinks) | 1.49 | (1.31-1.71) | <0.001 | 1.16 | (1.03-1.31) | 0.016 | 1.01 | (0.83-1.23) | 0.943 | 0.78 | (0.65-0.95) | 0.011 |
| Heavy Consumption (3+ drinks) | 0.83 | (0.73-0.95) | 0.007 | 0.90 | (0.79-1.02) | 0.086 | 0.84 | (0.69-1.01) | 0.064 | 0.90 | (0.75-1.09) | 0.273 |
| Physically Inactive | 0.58 | (0.42-0.79) | 0.001 | 0.88 | (0.69-1.13) | 0.324 | 1.00 | (0.63-1.59) | 0.992 | 1.53 | (1.00-2.33) | 0.050 |
| Activities of Daily Living | 0.98 | (0.97-0.99) | 0.002 | 1.00 | (0.99-1.00) | 0.309 | 0.99 | (0.97-1.00) | 0.032 | 1.00 | (0.98-1.01) | 0.590 |
| BMI | 1.20 | (1.12-1.28) | <0.001 | 1.11 | (1.04-1.19) | 0.002 | 0.92 | (0.86-0.99) | 0.024 | 0.85 | (0.79-0.92) | <0.001 |
| Hypertension | 1.15 | (1.00-1.33) | 0.046 | 1.05 | (0.92-1.18) | 0.486 | 1.12 | (0.92-1.37) | 0.264 | 1.02 | (0.84-1.23) | 0.865 |
| Heart Problem | 1.46 | (1.26-1.69) | <0.001 | 1.21 | (1.05-1.39) | 0.008 | 0.80 | (0.66-0.98) | 0.027 | 0.66 | (0.55-0.80) | <0.001 |
| Stroke | 1.27 | (1.01-1.59) | 0.042 | 1.12 | (0.89-1.40) | 0.326 | 0.79 | (0.60-1.04) | 0.096 | 0.70 | (0.52-0.92) | 0.011 |
| Diabetes | 1.14 | (0.98-1.32) | 0.089 | 1.17 | (1.02-1.34) | 0.026 | 0.85 | (0.69-1.04) | 0.111 | 0.87 | (0.72-1.06) | 0.168 |
| Cancer | 1.71 | (1.44-2.02) | <0.001 | 1.23 | (1.05-1.45) | 0.012 | 0.97 | (0.77-1.21) | 0.770 | 0.70 | (0.56-0.87) | 0.002 |
| Arthritis | 1.32 | (1.15-1.51) | <0.001 | 1.29 | (1.14-1.45) | <0.001 | 0.72 | (0.58-0.89) | 0.002 | 0.70 | (0.58-0.86) | 0.001 |
| Psychiatric Condition | 1.15 | (0.98-1.34) | 0.087 | 1.31 | (1.14-1.51) | <0.001 | 0.61 | (0.50-0.75) | <0.001 | 0.70 | (0.57-0.85) | <0.001 |

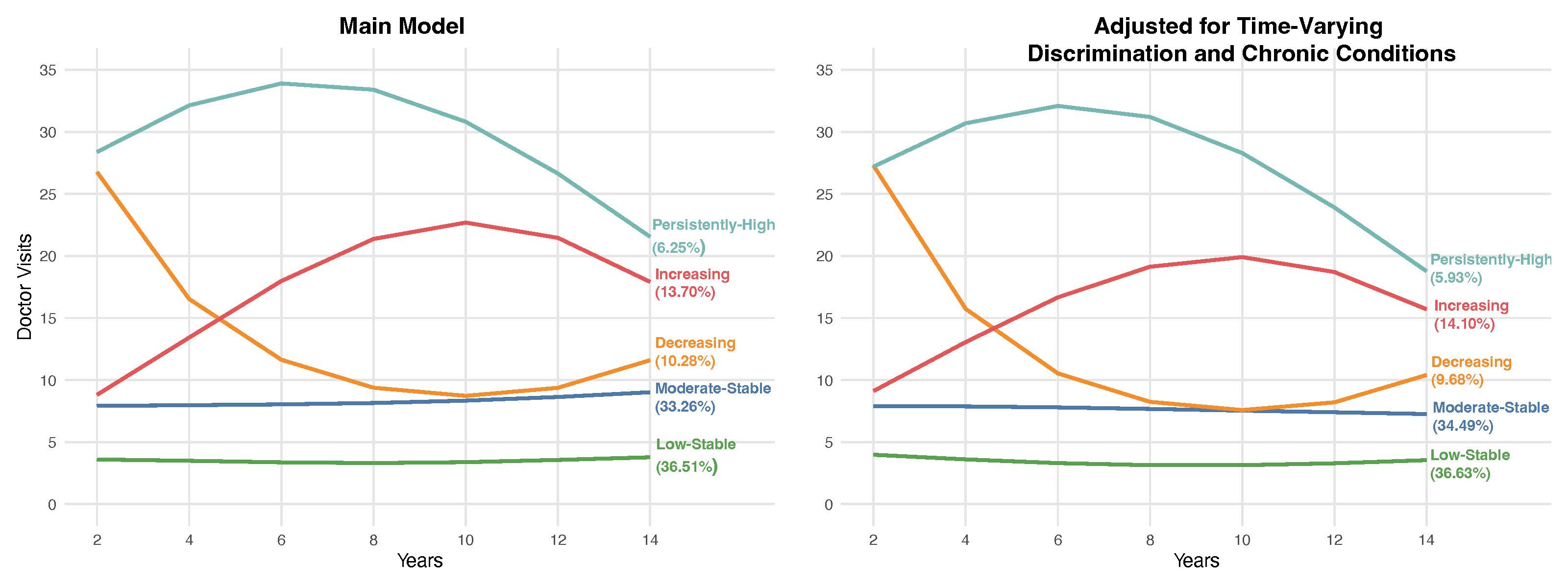

**Supplementary Figure 2.** Comparison of Estimated Trajectories of Doctor Visits Over Time in Middle-Aged and Older Adults, Health and Retirement Study 2008-2020. Note: The estimates were derived using a group-based trajectory model that assumes a zero-inflated Poisson distribution for the number of doctor visits over time*.*

| Trajectory Group | Moderate-Stable | | | Decreasing | | | Increasing | | | Persistently-High | | |
| --- | --- | --- | --- | --- | --- | --- | --- | --- | --- | --- | --- | --- |
| (Reference Group) | (Low-Stable) | | | (Low-Stable) | | | (Low-Stable) | | | (Low-Stable) | | |
|  | RRR | 95% CI | P-Value | RRR | 95% CI | P-Value | RRR | 95% CI | P-Value | RRR | 95% CI | P-Value |
| Unadjusted Model |  |  |  |  |  |  |  |  |  |  |  |  |
| Discrimination in Healthcare | 0.99 | (0.89-1.10) | 0.889 | 1.29 | (1.11-1.50) | 0.001 | 1.06 | (0.93-1.22) | 0.401 | 1.57 | (1.32-1.87) | <0.001 |
| Adjusted Model |  |  |  |  |  |  |  |  |  |  |  |  |
| Discrimination in Healthcare | 1.02 | (0.92-1.14) | 0.661 | 1.23 | (1.06-1.44) | 0.008 | 1.03 | (0.90-1.19) | 0.642 | 1.38 | (1.15-1.65) | 0.001 |
| Age | 1.02 | (1.01-1.02) | <0.001 | 1.02 | (1.01-1.03) | 0.001 | 1.02 | (1.01-1.02) | <0.001 | 1.01 | (1.00-1.02) | 0.257 |
| Male | 0.73 | (0.67-0.79) | <0.001 | 0.74 | (0.64-0.84) | <0.001 | 0.74 | (0.66-0.84) | <0.001 | 0.65 | (0.54-0.77) | <0.001 |
| Non-Hispanic Black | 0.82 | (0.73-0.92) | 0.001 | 0.80 | (0.67-0.95) | 0.010 | 0.75 | (0.64-0.87) | <0.001 | 0.67 | (0.53-0.83) | <0.001 |
| Hispanic | 0.80 | (0.67-0.95) | 0.009 | 0.99 | (0.77-1.27) | 0.950 | 0.90 | (0.72-1.12) | 0.349 | 0.66 | (0.46-0.93) | 0.018 |
| Married | 1.20 | (1.09-1.32) | <0.001 | 0.93 | (0.81-1.06) | 0.287 | 0.96 | (0.85-1.08) | 0.522 | 1.04 | (0.88-1.24) | 0.622 |
| Education | 1.08 | (1.07-1.10) | <0.001 | 1.07 | (1.04-1.10) | <0.001 | 1.08 | (1.06-1.11) | <0.001 | 1.1 | (1.06-1.13) | <0.001 |
| Wealth | 1.01 | (0.99-1.02) | 0.385 | 0.98 | (0.96-1.00) | 0.019 | 0.99 | (0.97-1.00) | 0.154 | 0.98 | (0.96-1.00) | 0.097 |
| Uninsured | 0.85 | (0.72-1.00) | 0.055 | 0.50 | (0.37-0.68) | <0.001 | 0.76 | (0.61-0.96) | 0.023 | 0.47 | (0.31-0.70) | <0.001 |
| Smoking Status |  |  |  |  |  |  |  |  |  |  |  |  |
| Former Smoker | 1.10 | (1.01-1.21) | 0.034 | 1.28 | (1.11-1.47) | 0.001 | 1.20 | (1.06-1.36) | 0.003 | 1.19 | (1.00-1.42) | 0.050 |
| Current Smoker | 0.93 | (0.82-1.06) | 0.258 | 1.15 | (0.95-1.39) | 0.159 | 1.09 | (0.92-1.30) | 0.296 | 1.19 | (0.94-1.50) | 0.153 |
| Drinking Status |  |  |  |  |  |  |  |  |  |  |  |  |
| No Consumption (0 drinks) | 1.03 | (0.93-1.15) | 0.535 | 1.44 | (1.22-1.71) | <0.001 | 1.20 | (1.04-1.38) | 0.014 | 1.65 | (1.33-2.06) | <0.001 |
| Heavy Consumption (3+ drinks) | 0.95 | (0.84-1.08) | 0.437 | 1.26 | (1.02-1.56) | 0.029 | 1.20 | (1.01-1.43) | 0.037 | 1.29 | (0.98-1.70) | 0.065 |
| Physically Inactive | 1.05 | (0.97-1.15) | 0.235 | 1.57 | (1.37-1.80) | <0.001 | 1.14 | (1.01-1.28) | 0.029 | 1.49 | (1.26-1.77) | <0.001 |
| Activities of Daily Living | 1.09 | (1.03-1.16) | 0.006 | 1.34 | (1.25-1.43) | <0.001 | 1.20 | (1.12-1.29) | <0.001 | 1.38 | (1.28-1.49) | <0.001 |
| BMI | 1.01 | (1.00-1.02) | 0.04 | 0.99 | (0.98-1.00) | 0.250 | 1.00 | (0.99-1.01) | 0.537 | 1.01 | (0.99-1.02) | 0.362 |
| Hypertension | 1.08 | (0.99-1.18) | 0.094 | 1.09 | (0.95-1.25) | 0.220 | 1.00 | (0.89-1.13) | 0.995 | 0.87 | (0.73-1.03) | 0.099 |
| Heart Problem | 0.95 | (0.85-1.06) | 0.341 | 1.14 | (0.97-1.33) | 0.121 | 1.05 | (0.90-1.21) | 0.544 | 1.29 | (1.07-1.57) | 0.009 |
| Stroke | 0.83 | (0.70-1.00) | 0.050 | 0.85 | (0.66-1.10) | 0.213 | 0.84 | (0.67-1.06) | 0.140 | 0.97 | (0.72-1.29) | 0.823 |
| Diabetes | 1.25 | (1.12-1.39) | <0.001 | 1.17 | (1.00-1.38) | 0.055 | 1.28 | (1.11-1.48) | 0.001 | 1.19 | (0.98-1.46) | 0.080 |
| Cancer | 0.92 | (0.81-1.05) | 0.238 | 1.42 | (1.19-1.70) | <0.001 | 1.01 | (0.85-1.19) | 0.934 | 1.30 | (1.04-1.61) | 0.021 |
| Arthritis | 1.02 | (0.93-1.11) | 0.654 | 1.18 | (1.03-1.35) | 0.017 | 1.26 | (1.12-1.42) | <0.001 | 1.50 | (1.26-1.79) | <0.001 |
| Psychiatric Condition | 1.08 | (0.96-1.21) | 0.212 | 1.02 | (0.86-1.21) | 0.815 | 1.29 | (1.12-1.50) | 0.001 | 1.58 | (1.31-1.91) | <0.001 |

**Supplementary Table 3**. Multinomial Estimates of Association between Discrimination in Healthcare Settings and Trajectory Group Membership including adjustment for Time-Varying Discrimination and Time-Varying Total Chronic Conditions, Health and Retirement Study

**Supplementary Table 4**. Multinomial Estimates of Association between Discrimination in Healthcare Settings and Trajectory Group Membership with varying Comparator Groups based on the Similar Number of Doctor Visits, including adjustment for Time-Varying Discrimination and Time-Varying Total Chronic Conditions, Health and Retirement Study

| Trajectory Group | Decreasing | | | Increasing | | | Decreasing | | | Increasing | | |
| --- | --- | --- | --- | --- | --- | --- | --- | --- | --- | --- | --- | --- |
| (Reference Group) | (Moderate-Stable) | | | (Moderate-Stable) | | | (Persistently-High) | | | (Persistently-High) | | |
|  | RRR | 95% CI | P-value | RRR | 95% CI | P-value | RRR | 95% CI | P-value | RRR | 95% CI | P-value |
| Unadjusted Model |  |  |  |  |  |  |  |  |  |  |  |  |
| Discrimination in Healthcare | 1.30 | (1.12-1.51) | 0.001 | 1.07 | (0.93-1.23) | 0.347 | 0.82 | (0.67-1.01) | 0.063 | 0.68 | (0.56-0.82) | <0.001 |
| Adjusted Model |  |  |  |  |  |  |  |  |  |  |  |  |
| Discrimination in Healthcare | 1.20 | (1.03-1.41) | 0.018 | 1.01 | (0.88-1.16) | 0.895 | 0.90 | (0.73-1.11) | 0.311 | 0.75 | (0.61-0.92) | 0.006 |
| Age | 1.00 | (0.99-1.01) | 0.938 | 1.00 | (0.99-1.01) | 0.965 | 1.01 | (1.00-1.02) | 0.159 | 1.01 | (1.00-1.02) | 0.130 |
| Male | 1.01 | (0.88-1.16) | 0.870 | 1.02 | (0.91-1.15) | 0.701 | 1.14 | (0.93-1.39) | 0.197 | 1.15 | (0.96-1.39) | 0.137 |
| Non-Hispanic Black | 0.97 | (0.82-1.16) | 0.749 | 0.91 | (0.78-1.06) | 0.229 | 1.2 | (0.93-1.54) | 0.163 | 1.12 | (0.88-1.43) | 0.363 |
| Hispanic | 1.24 | (0.96-1.60) | 0.095 | 1.13 | (0.90-1.42) | 0.311 | 1.51 | (1.02-2.24) | 0.039 | 1.37 | (0.94-2.00) | 0.103 |
| Married | 0.77 | (0.67-0.89) | <0.001 | 0.80 | (0.71-0.91) | <0.001 | 0.89 | (0.73-1.08) | 0.240 | 0.92 | (0.77-1.11) | 0.386 |
| Education | 0.99 | (0.96-1.01) | 0.304 | 1.00 | (0.98-1.02) | 0.929 | 0.97 | (0.94-1.01) | 0.160 | 0.99 | (0.95-1.02) | 0.433 |
| Wealth | 0.97 | (0.95-0.99) | 0.003 | 0.98 | (0.96-1.00) | 0.038 | 1.00 | (0.97-1.02) | 0.812 | 1.01 | (0.98-1.03) | 0.583 |
| Uninsured | 0.59 | (0.43-0.80) | 0.001 | 0.90 | (0.71-1.15) | 0.390 | 1.06 | (0.66-1.71) | 0.800 | 1.63 | (1.06-2.52) | 0.026 |
| Smoking Status |  |  |  |  |  |  |  |  |  |  |  |  |
| Former Smoker | 1.16 | (1.01-1.33) | 0.038 | 1.09 | (0.96-1.23) | 0.168 | 1.08 | (0.88-1.31) | 0.481 | 1.01 | (0.84-1.22) | 0.923 |
| Current Smoker | 1.24 | (1.02-1.51) | 0.033 | 1.18 | (0.99-1.40) | 0.062 | 0.97 | (0.74-1.27) | 0.82 | 0.92 | (0.71-1.19) | 0.544 |
| Drinking Status |  |  |  |  |  |  |  |  |  |  |  |  |
| No Consumption (0) | 1.40 | (1.18-1.66) | <0.001 | 1.16 | (1.00-1.34) | 0.045 | 0.87 | (0.67-1.13) | 0.299 | 0.72 | (0.57-0.92) | 0.008 |
| Heavy Consumption (3+) | 1.33 | (1.08-1.64) | 0.008 | 1.26 | (1.06-1.50) | 0.008 | 0.98 | (0.71-1.34) | 0.89 | 0.93 | (0.69-1.25) | 0.630 |
| Physically Inactive | 1.49 | (1.30-1.71) | <0.001 | 1.08 | (0.96-1.22) | 0.194 | 1.05 | (0.86-1.29) | 0.611 | 0.76 | (0.63-0.92) | 0.005 |
| Activities of Daily Living | 1.23 | (1.15-1.31) | <0.001 | 1.10 | (1.03-1.18) | 0.006 | 0.97 | (0.89-1.04) | 0.375 | 0.87 | (0.80-0.94) | <0.001 |
| BMI | 0.99 | (0.98-1.00) | 0.011 | 1.00 | (0.99-1.00) | 0.335 | 0.99 | (0.97-1.00) | 0.101 | 1.00 | (0.98-1.01) | 0.685 |
| Hypertension | 1.01 | (0.88-1.16) | 0.886 | 0.93 | (0.82-1.04) | 0.213 | 1.26 | (1.03-1.54) | 0.023 | 1.16 | (0.96-1.39) | 0.131 |
| Heart Problem | 1.20 | (1.02-1.41) | 0.026 | 1.10 | (0.96-1.28) | 0.177 | 0.88 | (0.70-1.09) | 0.246 | 0.81 | (0.66-1.00) | 0.048 |
| Stroke | 1.02 | (0.79-1.32) | 0.861 | 1.01 | (0.80-1.27) | 0.954 | 0.88 | (0.63-1.23) | 0.459 | 0.87 | (0.63-1.20) | 0.390 |
| Diabetes | 0.94 | (0.80-1.10) | 0.445 | 1.03 | (0.89-1.18) | 0.722 | 0.98 | (0.78-1.23) | 0.871 | 1.07 | (0.86-1.33) | 0.527 |
| Cancer | 1.54 | (1.29-1.84) | <0.001 | 1.09 | (0.92-1.29) | 0.316 | 1.10 | (0.86-1.40) | 0.458 | 0.78 | (0.61-0.99) | 0.042 |
| Arthritis | 1.16 | (1.01-1.33) | 0.037 | 1.24 | (1.10-1.39) | <0.001 | 0.79 | (0.64-0.96) | 0.020 | 0.84 | (0.70-1.02) | 0.078 |
| Psychiatric Condition | 0.95 | (0.80-1.12) | 0.53 | 1.20 | (1.04-1.39) | 0.015 | 0.65 | (0.52-0.81) | <0.001 | 0.82 | (0.67-1.01) | 0.058 |
